# West Nile Virus (*Orthoflavivirus nilense*) RNA concentrations in wastewater solids at five wastewater treatment plants in the United States

**DOI:** 10.1101/2025.04.07.25325420

**Authors:** Alessandro Zulli, Dorothea Duong, Bridgette Shelden, Amanda L. Bidwell, Marlene K. Wolfe, Bradley J. White, Alexandria B. Boehm

## Abstract

**Background:** *Orthoflavivirus nilense*, formerly known as West Nile Virus (WNV), has become endemic to the United States since its introduction in 1999. It has caused an estimated 4.2 million infections since then, and nearly 3,000 deaths. While 80% of infections remain asymptomatic, approximately half of confirmed cases develop into neuroinvasive disease, which is responsible for significant morbidity. Current surveillance methods rely primarily on mosquito pool testing, which is both costly and time-intensive, at best providing delayed epidemiological information. Wastewater-based epidemiology (WBE) has proven an effective method for the surveillance of various pathogens, including other orthoflaviviruses such as Dengue. Given current knowledge of WNV shedding, with over 50% of patients with neurological symptoms shedding it in urine, WBE represents a potentially valuable surveillance approach that has so far been underexplored.

**Methods:** This study evaluates the viability of using wastewater to assess community prevalence of WNV. We used a targeted droplet digital RT-PCR approach (ddRT-PCR) to measure WNV concentrations in wastewater retrospectively from five locations and in over 600 samples. Three of these locations were in communities with multiple confirmed WNV infections, while two were not. Samples were collected during periods corresponding to typical WNV seasonality (spring to fall). SARS-CoV-2 RNA was measured simultaneously to assess nucleic acid degradation during sample storage. Publicly available confirmed WNV case data were compiled from the California and Nebraska departments of public health and their weekly arboviral reports.

**Results:** We demonstrated that WNV RNA can be detected in wastewater samples during periods of known viral circulation within a community. We show that the adapted ddRT-PCR assay is highly specific and sensitive, and that detections in wastewater solids correspond to the occurrence of cases in the season and location of sampling. WNV was detected in 9 samples in 3 locations with known WNV clinical cases – wastewater positivity rates in these locations ranged from 3.3% to 13%. The results suggest that wastewater monitoring could serve as an effective complement to traditional surveillance methods, particularly for sentinel surveillance in locations which do not have extensive mosquito and clinical testing systems.

## Introduction

*Orthoflavivirus nilense*, formerly West Nile Virus (WNV), is a mosquito-borne single-stranded RNA virus first identified in Uganda in 1937 that has since been introduced to every continent except Antarctica (Chancey et al., 2015). The virus is spread in humans by the *Culex* mosquito, which acts as the transmission vector between birds and humans (Chancey et al., 2015; World Health Organization). Since Centers for Disease Control (CDC) surveillance of the virus began in 1999, the virus has caused an estimated 4.2 million infections, 59,141 confirmed cases, 27,617 hospitalizations, and 2,958 deaths (CDC, 2025). While an estimated 80% of cases are asymptomatic, approximately half of confirmed cases were neuroinvasive, often causing significant morbidity and early disability (McDonald, 2021; Santini et al., 2022; CDC, 2025). The impact of the virus in the United States is therefore significant, and the growing ecological range of the *Culex* mosquito due to climate change has led to West Nile virus becoming a significant public health concern (Paz, 2015; Heidecke, Schettini & Rocklöv, 2023; Erazo et al., 2024).

Since its introduction in 1999, WNV has become endemic to the United States, consistently causing thousands of human cases each year, along with sporadic epizootics and avian/equine infections (Ronca, Ruff & Murray, 2021; CDC, 2025). Typically, human cases peak in late summer and early fall, and then rapidly decrease as temperatures drop below 18°C, at which point the virus rarely establishes itself in *Culex* mosquitoes (Di Pol, Crotta & Taylor, 2022; Vollans et al., 2024). As there is no human vaccine available, surveillance and control methods primarily focus on mosquito pool surveillance, mosquito control, and public health education (Ronca, Ruff & Murray, 2021). Mosquito control includes various strategies, such as larval control, adult mosquito control, and public health surveillance. Many states, such as Florida and Nevada, regularly test mosquito pools for arboviruses during spring, summer and fall, allowing for granular mapping of the virus’ prevalence.(Florida Department of Health, 2025; Southern Nevada Health District, 2025) However, these methods are both costly and time-intensive, requiring dedicated teams and systems, and often providing epidemiological information with a significant delay.

Wastewater based epidemiology (WBE) has demonstrated to be an effective method for tracking and providing epidemiological information on various human pathogens including SARS-CoV-2, influenza, Chikungunya and Dengue virus (Peccia et al., 2020; Boehm et al., 2023a; Monteiro et al., 2023; Wolfe et al., 2024). In patients with WNV infections demonstrating neurological symptoms, over 50% of urine samples were found to be positive for WNV and at higher relative concentrations than cerebrospinal fluid – 50.5% of all urine samples compared to 2.8% of cerebrospinal fluid samples – indicating that the virus is likely shed into wastewater streams (Gdoura et al., 2022). Note that the authors did not provide concentrations in externally valid units; they provided cycle threshold (CT) values from a quantitative PCR instrument. Further, studies have shown that WNV preferentially partitions into wastewater solids (Roldan-Hernandez, Van Oost & Boehm, 2024), which are typically used for WBE. Given the wastewater detections of other flaviviruses such Dengue virus, known shedding of WNV in the urine of infected patients, and the difficulty in implementing environmental and clinical surveillance systems for WNV, WBE could provide a rapid, cost-effective method for gathering epidemiological information on entire communities (Gyure, 2009; Barzon et al., 2013; Gdoura et al., 2022; Wolfe et al., 2024). Despite the potential advantages of WBE for arboviral surveillance, there has been limited investigation into its application for WNV monitoring.

In this study, we assess the viability of using wastewater to assess community infections of WNV. We use a targeted droplet digital RT-PCR approach to measure WNV concentrations in over 600 wastewater samples from five different wastewater treatment plants. These results demonstrate the potential promise and limitations of WBE for sentinel monitoring of the spread of WNV.

## Methods

### WNV assay

We used a previously published hydrolysis-probe assay targeting the polyprotein gene of WNV (Lanciotti et al., 2000) in droplet digital RT-PCR (Table 1). The assay was tested in vitro for specificity and sensitivity using a virus panels (NATtrol Respiratory Verification Panel NATRVP2.1-BIO), and synthetic target nucleic acids (VR-3274SD) purchased from American Type Culture Collection (ATCC, Manassas, VA), respectively. The respiratory virus panel includes chemically inactivated intact influenza viruses, parainfluenza viruses, adenovirus, rhinovirus A, metapneumovirus, rhinovirus, RSV, several coronaviruses, and SARS-CoV-2. Nucleic acids were extracted from intact viruses using Chemagic Viral DNA/RNA 300 Kit H96 for Chemagic 360 (PerkinElmer, Waltham, MA). The assay was tested *in silico* for specificity by blasting the sequences in National Center for Biotechnology Information (NCBI) in October 2023. We first ran an inclusionary BLAST to confirm it matched recent WNV genomes, and then an exclusionary BLAST search excluding all WNV genomes, thereby only giving off-target matches

**Table 1.**
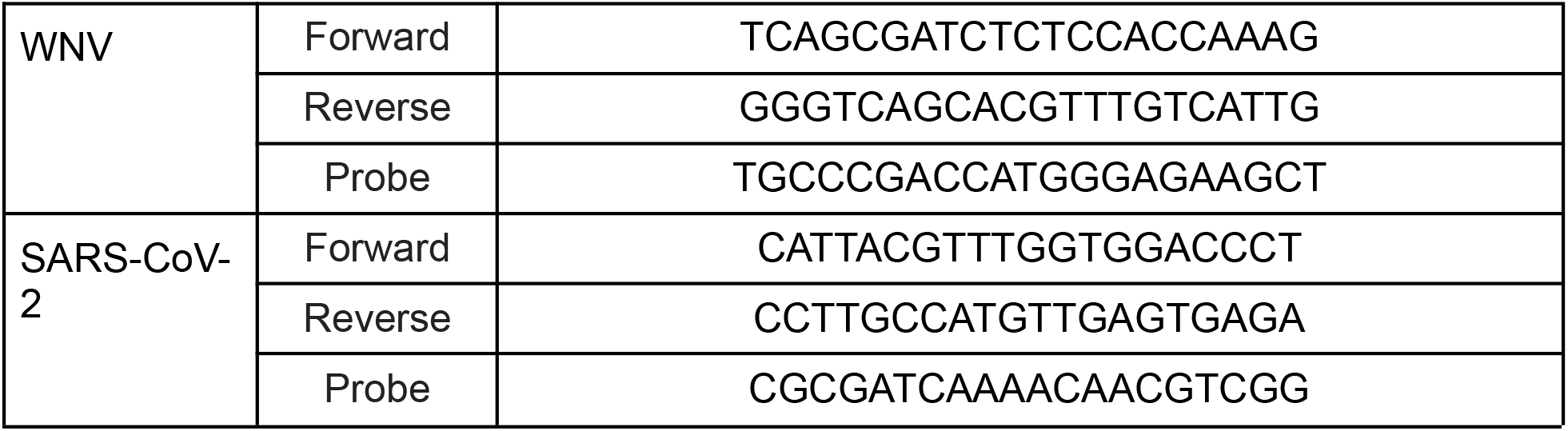
Primers and probes used in this study.

For in vitro testing, nucleic acids were used undiluted as template in digital droplet (RT-)PCR singleton assays for sensitivity and specificity testing in single wells. The concentration of targets used in the in vitro specificity testing was between 10^3^ and 10^4^ copies per well. Negative RT-PCR controls were included on each plate.

### Wastewater analyses

Wastewater samples from five different wastewater treatment plants (WWTPs) were tested for WNV RNA (Figure 1). Two WWTPs, San Jose (SJ) and Oceanside (OSP), are located in the San Francisco Bay Area of California, USA and serve 1,500,000 and 250,000 people, respectively. We processed approximately two samples per week over a 26.5 month period (2/1/21 - 4/14/23, month/day/year format, n = 229 for SJ and n = 230 for OSP). SJ and OSP sites were chosen to represent sites where contributing communities were not expected to have WNV infections based on available case data (California Department of Public Health, 2025). Two WWTPs, Sacramento (SAC) and Woodland (WD), are located within or adjacent to Sacramento County, CA, USA and serve 1,480,000 and 59,000 people, respectively. Samples were collected approximately three times per week between 6/2/23 and 10/20/23 at SAC (n = 60) and between 6/2/23 and 10/11/23 at WD (n = 51). One WWTP was located in Lincoln, NE (NE) and serves 240,000 people; three samples were collected per week between 8/2/23 and 10/11/23 (n = 31). SAC, WD, and NE sites were included in the study because they had multiple confirmed WNV infections in the contributing populations during the time of sample collection (Nebraska Department of Health and Human Services, 2025). Additional details of the sites can be found elsewhere (Boehm et al., 2024).

**Figure 1.**
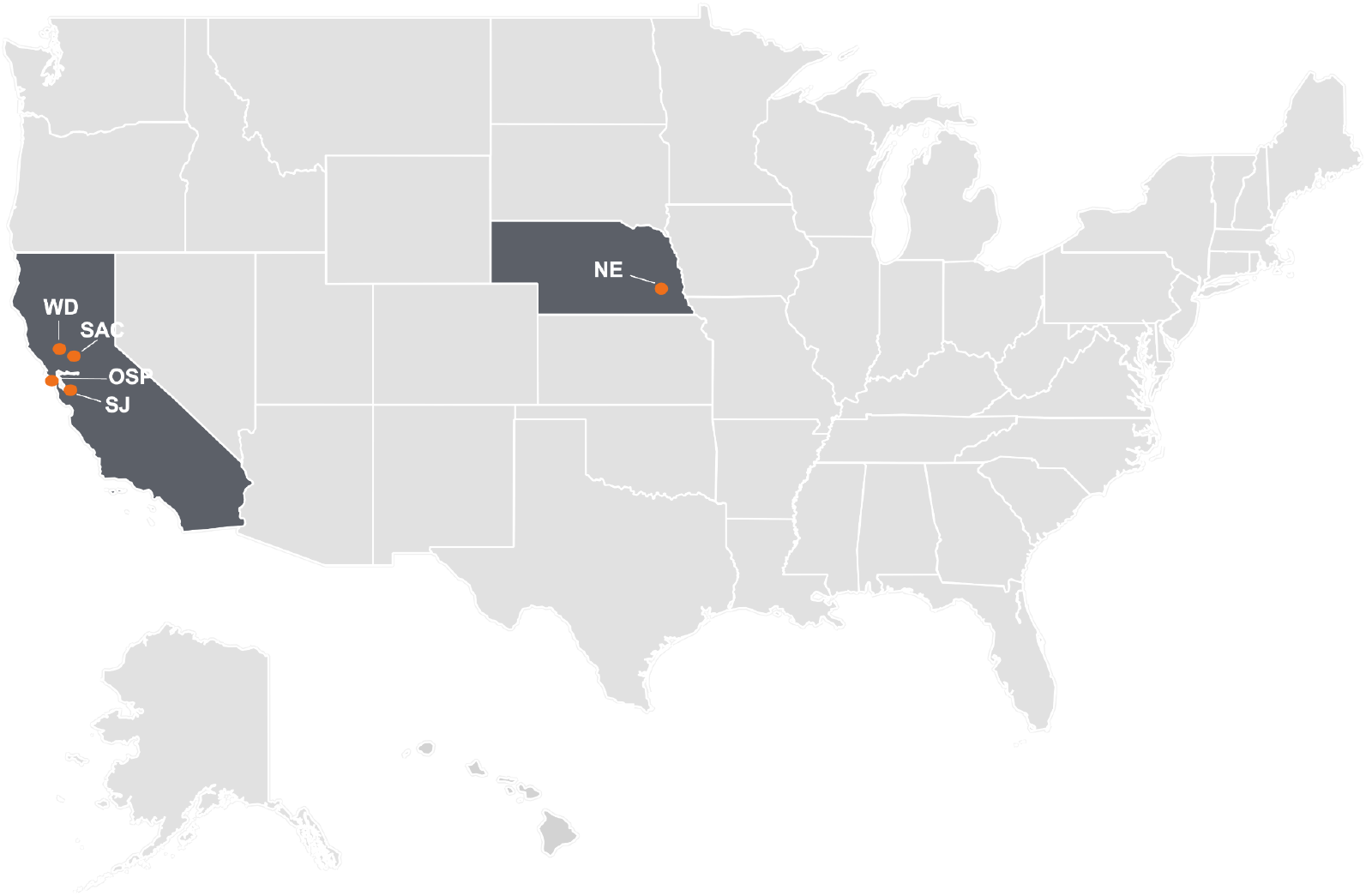
Map of the wastewater treatment plants (WWTPs) enrolled in this study. The orange dots represent the location of the five WWTPs and states with participating WWTPs are shaded in dark gray. This figure was generated using ArcGIS desktop; map layer from The United States Census Bureau’s Cartographic Boundary Files (https://www.census.gov/geographies/mapping-files/time-series/geo/carto-boundary-file.html)

At SJ, SAC, and OSP, 50 mL of settled solids from the primary clarifier, and at WD and NE, 50 mL of raw influent sewage were collected; sterile technique and clean bottles were used in all cases. Samples were stored at 4°C, transported to the lab, and processed within 48 hours of collection. Wastewater solids were isolated from the raw influent using centrifugation, as described previously (Boehm et al., 2024). Then settled solids from all samples were dewatered (Topol et al., 2021a). At this point in processing (prior to nucleic-acid extraction), samples from SJ and OSP were frozen at -80°C for 4 - 60 weeks, and then thawed overnight at 4°C prior to further processing. Remaining samples were not frozen before nucleic-acid extraction.

Nucleic acids were obtained from the dewatered solids following previously published protocols (Topol et al., 2021b; Boehm et al., 2023b). Those protocols include suspending the solids in a buffer at a concentration of 75 mg/ml and using an inhibitor removal kit; together these processes alleviate potential inhibition while maintaining good assay sensitivity (Huisman et al., 2022; Boehm et al., 2023b). Nucleic-acids were obtained from 6 or 10 replicate sample aliquots (6 replicates for SAC, NE, and WD; and 10 replicates for OSP and SJ). Each replicate nucleic-acid extract from each sample was subsequently stored between 8 and 273 days (median 266 days) for SJ, 1 and 8 days (median 5 days) for OSP, 14 and 154 days (median 84 days) for SAC, 32 and 163 days (median 98 days) for WD, and 32 and 102 days (median 67 days) for NE at -80°C and subjected to a single freeze thaw cycle. Upon thawing, WNV and SARS-CoV-2 RNA were measured immediately using multiplex digital droplet RT-PCR. SARS-CoV-2 RNA was used as a storage control as it was measured previously in the samples without any storage (methods and results reported in a Data Descriptor (Boehm et al., 2024).

The WNV and SARS-CoV-2 N gene assay were run in multiplex using a probe-mixing approach and unique fluorescent molecules (HEX, FAM, Cy5, Cy5.5, ROX, and/or ATTO950). Nucleic-acid targets from from SJ and OSP were measured in multiplex for SARS-CoV-2 (fluorescent molecule(s) on probe: FAM/HEX) and WNV RNA (ROX) along with assays for 3 other human viral targets not reported herein (rotavirus (FAM), human adenovirus group F (ROX/ATTO590), human norovirus GII (ATTO590)). Nucleic-acid targets from SAC, WD, and NE samples were measured in a duplex assay for SARS-CoV-2 (HEX) and WNV (FAM) RNA. Each nucleic acid extract was run in a single well so that 6 or 10 replicate wells, depending on the site, were run for each sample for each assay.

Each 96-well PCR plate of wastewater samples included PCR positive controls for each target assayed on the plate in 1 well, PCR negative no template controls in two wells, and extraction negative controls (consisting of water and lysis buffer) in two wells. PCR positive controls consisted of synthetic viral WNV gRNA (VR-3274SD, ATCC) and SARS-CoV-2 gRNA (VR-1986D, ATCC).

ddRT-PCR was performed on 20 µl samples from a 22 µl reaction volume, prepared using 5.5 µl template, mixed with 5.5 µl of One-Step RT-ddPCR Advanced Kit for Probes (Bio-Rad 1863021), 2.2 µl of 200 U/µl Reverse Transcriptase, 1.1 µl of 300 mM dithiothreitol (DDT) and primers and probes mixtures at a final concentration of 900 nM and 250 nM respectively. Primer and probes for assays were purchased from Integrated DNA Technologies (IDT, San Diego, CA) (Table 2). WNV and SARS-CoV-2 RNA was measured in reactions with undiluted template. Primers and probes are provided in Table 1.

Droplets were generated using the AutoDG Automated Droplet Generator (Bio-Rad, Hercules, CA). PCR was performed using Mastercycler Pro (Eppendforf, Enfield, CT) with with the following cycling conditions: reverse transcription at 50°C for 60 minutes, enzyme activation at 95°C for 5 minutes, 40 cycles of denaturation at 95°C for 30 seconds and annealing and extension at 59°C (for WNV and SARS-CoV-2) for 30 seconds, enzyme deactivation at 98°C for 10 minutes then an indefinite hold at 4°C. The ramp rate for temperature changes were set to 2°C/second and the final hold at 4°C was performed for a minimum of 30 minutes to allow the droplets to stabilize.

Droplets were analyzed using the QX200 or the QX600 Droplet Reader (Bio-Rad). A well had to have over 10,000 droplets for inclusion in the analysis. All liquid transfers were performed using the Agilent Bravo (Agilent Technologies, Santa Clara, CA).

Thresholding was done using QuantaSoft™ Analysis Pro Software (Bio-Rad, version 1.0.596) and QX Manager Software (Bio-Rad, version 2.0). Replicate wells were merged for analysis of each sample. In order for a sample to be recorded as positive, it had to have at least 3 positive droplets.

Concentrations of RNA targets were converted to concentrations in units of copies (cp)/g dry weight using dimensional analysis. The total error is reported as standard deviations and includes the errors associated with the Poisson distribution and the variability among the 6 or 10 replicates. Three positive droplets across 6 to 10 merged wells corresponds to a concentration between ∼500-1000 cp/g; the range in values is a result of the range in the equivalent mass of dry solids added to the wells. Wastewater measurements are available from the Stanford Digital Repository (https://purl.stanford.edu/pp102gy1970).

### Case data

For the SJ, OSP, SAC, and WD locations, clinical case data was compiled at the county level from the Vector-Borne Disease Section of the California Department of Public Health, which publishes weekly data on detections in humans and animals of West Nile Virus and St. Louis encephalitis virus (California Department of Public Health, 2025). For the NE location, clinical case data for Nebraska was compiled from the Nebraska Department of Health and Human Services, which publishes weekly mosquito borne disease reports that include WNV for the entire state (Nebraska Department of Health and Human Services, 2025). Clinical case data was collected for the same time periods as wastewater surveillance (SJ, OSP: 2/1/21 - 4/14/23, WD, SAC: 6/2/23 - 10/20/23, NE: 8/2/23 - 10/11/23). All case data are publicly available.

### Statistics

Case data were normalized by the populations of the counties or states included in the case reporting to calculate incidence rates. As case data are available for morbidity and mortality weekly report (MMWR) weeks, we created variables to indicate whether WNV RNA was detected each MMWR week for each WWTP. We calculated Kendall’s tau between WNV RNA detection each week (a binary variable) and the incidence rate of WNV infections in the contributing community to test the null hypothesis that there is no association. We also combined all the WWTP WNV RNA and incidence rate data together into one data set to test the same null hypothesis. Finally, we calculated the positivity rate across all samples from each of the five WWTPs, and the incidence rate of WNV infections across the entire time period each WWTP was studied and tested the null hypothesis that there is no correlation across the two variables. We used Kendall’s tau because the independent variables are not normally distributed. We used a p value less than 0.05 to reject the null hypothesis.

## Results

### WNV assay specificity and sensitivity

We used a previously published assay for WNV (Lanciotti et al., 2000) and that study conducted thorough sensitivity and specificity assessments. In order to ensure that the assay remained sensitive and specific, we carried out both i*n silico* and *in vitro* analyses. Both analyses indicated that the WNV assay was specific and sensitive. The assay matched downloaded WNV sequences from a global database (NCBI), and there was no cross reactivity with non-target sequences *in vitro*.

### QA/QC

All positive and negative controls were positive and negative, respectively. In a previous study, we provide RNA recovery from the samples, as inferred from recovery of a spiked bovine coronavirus, to be close to 1 (Boehm et al., 2024), and those results are not repeated herein. Median ratio of SARS-CoV-2 N gene measurements made in this study to those made using fresh samples was 0.2 at SJ and 0.4 at OSP suggesting the storage of wastewater solids and subsequent freeze thaw may have reduced measurement concentrations, but by less than an order of magnitude. Median ratio was 1.5 at NE, 1.6 at WD, and 1.3 at SAC suggesting that the storage of nucleic-acids did not reduce quantification and may have slightly enhanced it. Recall that storage of samples from the latter three WWTPs just involved storage of nucleic-acids, whereas storage of samples from SJ and OSP involved storage of wastewater solids and nucleic-acids. Although not ideal, storage of samples is essential for retrospective work like this. Additional QA/QC details and reporting that follows those recommend by the Environmental Microbiology Minimal Information (EMMI) guidelines (Borchardt et al., 2021) are provided at the Stanford Digital Repository (https://purl.stanford.edu/pp102gy1970).

### WNV in wastewater

We measured WNV RNA in 601 samples across five WWTPs. The vast majority (n=592/601, 98%) of the samples were negative for WNV RNA meaning the concentration was less than the lowest measurable concentration (approximately 1000 cp/g). Nine samples (2%) were positive for WNV RNA with concentrations ranging from 1755 cp/g to 11404 cp/g (median = 7146 cp/g). Positive samples were only observed at three of the sites: SAC (n=2 positive samples), WD (n = 3 positive samples), and NE (n=4 positive samples). No samples were positive at SJ or OSP. Positivity rates (number positive / total samples run) for SAC, WD, and NE are 3.3%, 5.9%, and 13%, respectively.

### Confirmed WNV cases

Weekly WNV case data are available aggregated across each county for California. In SAC, WD, SJ, and OSP, there were 29, 22, 1, and 1 cases respectively in the corresponding counties. Normalized by the county population, that is an incidence rate of 18, 100, 1, and 1 out of one million people for SAC, WD, SJ and OSP, respectively. Weekly WNV case data are available aggregated at the state for NE. There were 134 cases across the state, or an incidence rate of 67 out of one million people. Data for SAC, WD, and NE are provided in Figure 2; cases for SJ and OSP were recorded for the week of 10/7/22 and 10/29/21, respectively.

**Figure 2.**
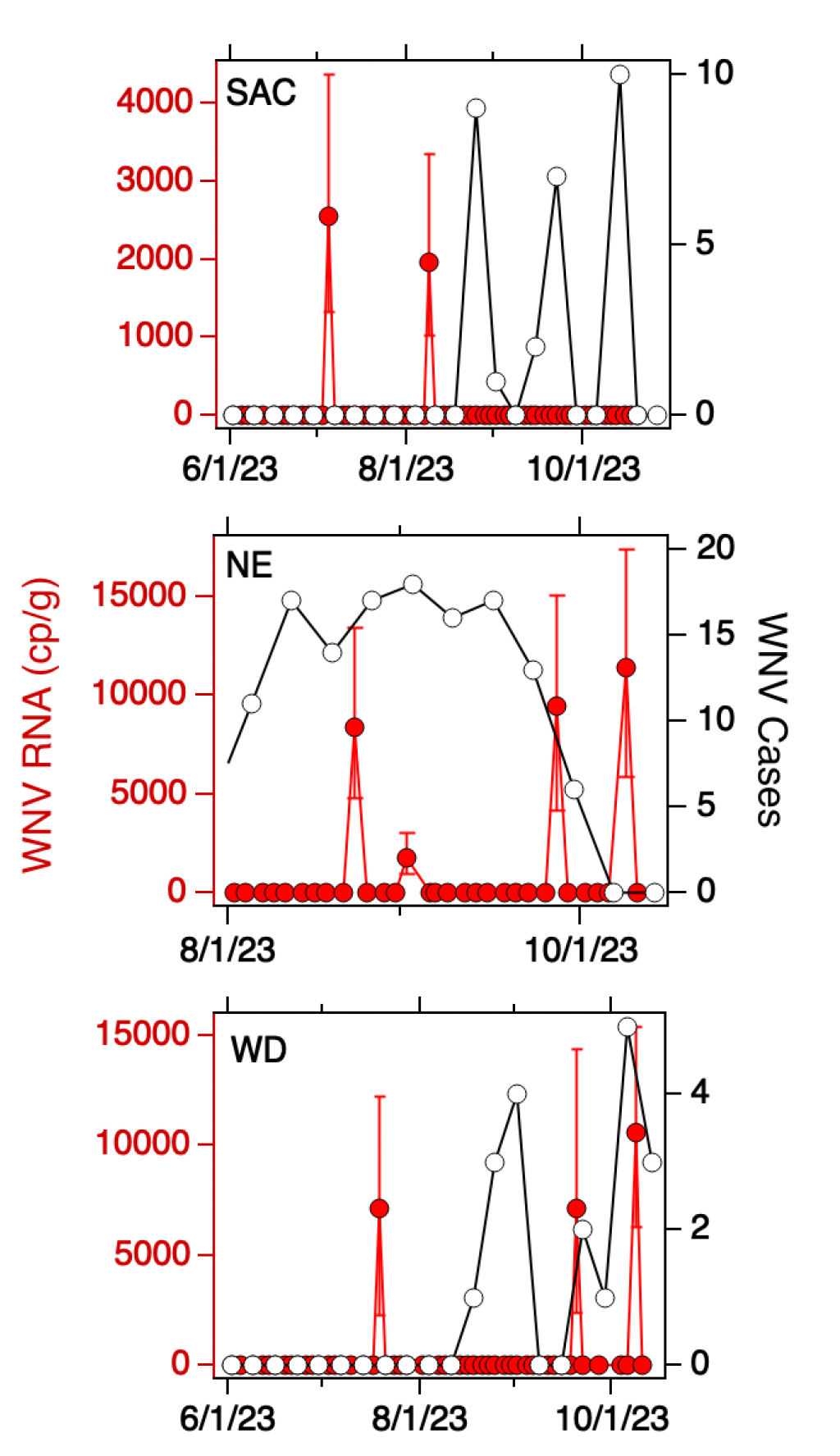
Concentrations of WNV RNA measured at SAC, NE, and WD sites during the study (in red, left axis) and confirmed cases of WNV infection in the associated county (SAC and WD) or state (NE) (in black, open circles, right axis). Error bars on the WNV RNA measurements represent standard deviations. A 0 value for concentration was imputed for samples where WNV RNA was not detected.

### Relationship between WNV RNA in wastewater and WNV cases

For each of the three WWTPs where WNV RNA was detected, there is no association between the weekly presence of WNV RNA and the WNV infection incidence rate (tau = -0.17, 0.20, and 0.24; p = 0.42, 0.34, and 0.42; n = 21, 21, and 10 for SAC, WD, and NE, respectively). When data from all five sites were combined, there is a significant association between weekly presence of WNV RNA and incidence rate (tau = 0.33, p = 2.6 x 10^-8^, n = 282). The wastewater positivity rate and county or state-specific (for NE only) incidence rate, over the entire duration of the study, were positively correlated, but the correlation was not statistically significant (tau = 0.74, p = 0.077, n=5).

## Discussion

WNV RNA was rarely detected in wastewater solids of three of the five WWTPs in this study. Detection of the viral RNA was not expected, and not observed, in two of the WWTPs where the most samples were analyzed; these WWTPs are located in a highly urbanized area in the San Francisco Bay Area of California where WNV infections are rare, and mosquito control programs are in place to control mosquito populations. On the other hand, WNV was detected at the three other WWTPs, with detection rates ranging from 3.3% (SAC) to 13% (NE). These three WWTPs were located in areas where WNV infections occur, and during times when WNV infections were recorded.

Although within each WWTP there is no association between detection of WNV RNA and incident cases of WNV, an aggregated analysis demonstrated that weekly presence of WNV RNA was significantly correlated to incidence rate. Therefore, detection of WNV RNA is suggestive of WNV cases in the community contributing to the WWTP during that WNV season, and could potentially be used as a supplementary or low-cost method for monitoring WNV in low resource settings and the increasing ecological range of WNV (Paz, 2015; Di Pol, Crotta & Taylor, 2022; Heidecke, Schettini & Rocklöv, 2023). This finding adds to the existing evidence that WBE may be a useful tool in tracking and combating the spread of arboviruses.

Previous implementations of WBE for arbovirus detection have yielded similar results to the ones presented in this study, showing sporadic detections of viruses in endemic areas and during acute epidemics (Monteiro et al., 2023, 2025; Fanok et al., 2023; Wolfe et al., 2024). These viruses include Dengue, Chikungunya, and Japanese Encephalitis, with each showing sporadic detection patterns during outbreaks in the underlying community. Wolfe et al. (ref) demonstrated that even in areas with very low prevalence of Dengue (Miami), detection was feasible, with 21% of samples positive for Dengue during a period with known clinical cases. Similarly, Monteiro et al. showed that in Portugal, where rates of Dengue and Chikungunya are low, Dengue was detected in 25% of samples, and Chikungunya in 11%. These demonstrate that arboviruses, and specifically flaviviruses (which include Dengue and WNV), are detectable through WBE in a manner consistent with underlying human transmission. Fanok et al. then showed that during an outbreak of Japanese Encephalitis Virus (JEV), 33% of samples tested had detectable levels of the virus (Fanok et al., 2023). This finding is particularly relevant as JEV is a flavivirus closely related to WNV for which humans are also dead-end hosts. Finally, a pre-print by Kuhn et al. showed that WNV is sporadically detected in wastewater in Oklahoma, reinforcing this pattern (Kuhn et al., 2024). More specifically, Kuhn et al. detected WNV in four WWTPs across three counties, though no significant correlation to clinical or animal cases was found, leaving open the question as to whether detections of WNV in wastewater reflect the underlying human epidemiological situation. Our results now demonstrate that these detections are not only possible and seasonal, but that the presence of WNV in wastewater is indicative of clinical cases in humans, an important step in the implementation process of WBE for WNV.

WBE for arboviruses can be an important tool in combating the spread of arboviruses, particularly as climate change is rapidly changing and expanding the ecological ranges of the vectors that carry arboviral disease, including the *Culex* mosquitoes responsible for WNV (Gilbert, 2021; Heidecke, Schettini & Rocklöv, 2023; Erazo et al., 2024). As average temperatures rise in certain locations, many will cross the 18 °C threshold at which WNV establishes itself in mosquitoes. These increases in temperature will also lead to higher rates of human transmission as vectors and their feeding patterns increase (Paz, 2015). This means that geographic areas that do not regularly see endemic transmission of WNV virus will be faced with the challenge of rapidly establishing clinical and environmental systems for control such as the ones present in California, Florida, and Nebraska (California Department of Public Health, 2025; Florida Department of Health, 2025; Nebraska Department of Health and Human Services, 2025). These systems are capital and time-intensive, but WBE provides a potential alternative — a low-cost, sentinel surveillance system that can build upon an established network for monitoring other viruses such as SARS-CoV-2 and influenza. Our results demonstrate that WNV WBE is a relatively simple addition to existing systems that can provide real-time information on the spread of these viruses in human populations, allowing public health officials to react accordingly.

There are limitations associated with this work. The available case data are limited in that they rely on confirmed, symptomatic, and likely severe WNV infections with neurological complications. WNV infections can be mild or asymptomatic, and such cases would not be counted by the current health surveillance systems (Santini et al., 2022; World Health Organization). There are limited data on WNV RNA shedding via human excretions, particularly given the context that humans are a dead end host for WNV. If shedding is low, then concentrations expected in wastewater will also be low. Although wastewater solids can provide sensitive detection of rare nucleic-acid targets owing to their natural ability to concentrate viral biomarkers, including WNV RNA, via adsorption (Roldan-Hernandez, Van Oost & Boehm, 2024), perhaps an even more sensitive approach will be needed to measure expected low levels of WNV RNA in wastewater.

## Conclusion

This study demonstrates that WNV RNA can be detected in wastewater samples using a targeted ddRT-PCR approach, and that these detections correlate with known cases of WNV infection across the communities sampled. While detections were sporadic, ranging from 3.3% to 13%, they were exclusively found in areas with established WNV transmission (SAC, WD, and NE) and absent in locations with minimal WNV activity (SJ and OSP). The significant correlations between weekly WNV RNA presence and incidence rates in the aggregated analysis suggests that wastewater surveillance has potential as a complementary tool to traditional WNV monitoring systems such as clinical, animal, and mosquito pool surveillance.

As climate change expands the ecological range of the Culex mosquito vector and increases the risk of WNV transmission in new geographic areas, wastewater-based epidemiology offers a cost-effective surveillance approach that can leverage pre-existing infrastructure. While challenges remain, including limited understanding of viral shedding from humans as dead end hosts, this study provides important evidence that wastewater-based epidemiology can serve as a viable sentinel surveillance system for WNV. By integrating this approach into existing WNV monitoring strategies, public health officials may gain additional context necessary to implement targeted interventions and mitigate the impact of WNV in both historically and newly endemic communities.

## Data Availability

All data produced are available online at the Stanford Digital Repository https://purl.stanford.edu/pp102gy1970

## Acknowledgments

We thank the participating wastewater treatment plants for their samples for the project.

## References

Barzon L, Pacenti M, Franchin E, Pagni S, Martello T, Cattai M, Cusinato R, Palù G. 2013. Excretion of West Nile Virus in Urine During Acute Infection. The Journal of Infectious Diseases 208:1086–1092. DOI: 10.1093/infdis/jit290.

Boehm AB, Hughes B, Duong D, Chan-Herur V, Buchman A, Wolfe MK, White BJ. 2023a. Wastewater concentrations of human influenza, metapneumovirus, parainfluenza, respiratory syncytial virus, rhinovirus, and seasonal coronavirus nucleic-acids during the COVID-19 pandemic: a surveillance study. The Lancet Microbe 4. DOI: 10.1016/S2666-5247(22)00386-X.

Boehm AB, Wolfe MK, Bidwell AL, Zulli A, Chan-Herur V, White BJ, Shelden B, Duong D. 2024. Human pathogen nucleic acids in wastewater solids from 191 wastewater treatment plants in the United States. Scientific Data 11:1141. DOI: 10.1038/s41597-024-03969-8.

Boehm AB, Wolfe MK, Wigginton KR, Bidwell A, White BJ, Hughes B, Duong, Chan-Herur V, Bischel HN, Naughton CC.2023b. Human viral nucleic acids concentrations in wastewater solids from Central and Coastal California USA. Scientific Data 10:396.

Borchardt MA, Boehm AB, Salit M, Spencer SK, Wigginton KR, Noble RT. 2021. The Environmental Microbiology Minimum Information (EMMI) Guidelines: qPCR and dPCR Quality and Reporting for Environmental Microbiology. Environmental Science & Technology 55:10210–10223. DOI: 10.1021/acs.est.1c01767.

California Department of Public Health. 2025. Westnile.ca.gov | California West Nile Virus Website. Available at https://westnile.ca.gov/faq (accessed April 1, 2025).

CDC. 2025. Historic Data (1999-2023). Available at https://www.cdc.gov/west-nile-virus/data-maps/historic-data.html (accessed March 30, 2025).

Chancey C, Grinev A, Volkova E, Rios M. 2015. The Global Ecology and Epidemiology of West Nile Virus. BioMed Research International 2015:376230. DOI: 10.1155/2015/376230.

Di Pol G, Crotta M, Taylor RA. 2022. Modelling the temperature suitability for the risk of West Nile Virus establishment in European Culex pipiens populations. Transboundary and Emerging Diseases 69:e1787–e1799. DOI: 10.1111/tbed.14513.

Equine Arbovirus Dashboard. Available at https://www.aphis.usda.gov/livestock-poultry-disease/equine/arbovirus-dashboard (accessed March 30, 2025).

Erazo D, Grant L, Ghisbain G, Marini G, Colón-González FJ, Wint W, Rizzoli A, Van Bortel W, Vogels CBF, Grubaugh ND, Mengel M, Frieler K, Thiery W, Dellicour S. 2024. Contribution of climate change to the spatial expansion of West Nile virus in Europe. Nature Communications 15:1196. DOI: 10.1038/s41467-024-45290-3.

Fanok S, Monis PT, Keegan AR, King BJ. 2023. The detection of Japanese encephalitis virus in municipal wastewater during an acute disease outbreak. Journal of Applied Microbiology 134:lxad275. DOI: 10.1093/jambio/lxad275.

Florida Department of Health. 2025. Mosquito-Borne Disease Surveillance | Florida Department of Health. Available at https://www.floridahealth.gov/diseases-and-conditions/mosquito-borne-diseases/surveillance.html (accessed March 30, 2025).

Gdoura M, Fares W, Bougatef S, Inoubli A, Touzi H, Hogga N, Ben Dhifallah I, Hannachi N, Argoubi A, Kacem S, Karray H, Ben Alaya N, Triki H. 2022. The value of West Nile virus RNA detection by real-time RT-PCR in urine samples from patients with neuroinvasive forms. Archives of Microbiology 204:238. DOI: 10.1007/s00203-022-02829-6.

Gilbert L. 2021. The Impacts of Climate Change on Ticks and Tick-Borne Disease Risk. Annual Review of Entomology 66:373–388. DOI: 10.1146/annurev-ento-052720-094533.

Gyure KA. 2009. West Nile Virus Infections. Journal of Neuropathology & Experimental Neurology 68:1053–1060. DOI: 10.1097/NEN.0b013e3181b88114.

Heidecke J, Schettini AL, Rocklöv J. 2023. West Nile virus eco-epidemiology and climate change. PLOS Climate 2:e0000129. DOI: 10.1371/journal.pclm.0000129.

Huisman JS, Scire J, Caduff L, Fernandez-Cassi X, Ganesanandamoorthy P, Kull A, Scheidegger A, Stachler E, Boehm AB, Hughes B, Knudson A, Topol A, Wigginton KR, Wolfe MK, Kohn T, Ort C, Stadler T, Julian TR. 2022. Wastewater-based estimation of the effective reproductive number of SARS-CoV-2. Environmental Health Perspectives 130:057011–1.

Kuhn KG, Shelton K, Sanchez GJ, Zamor RM, Bohanan K, Nichols M, Morris L, Robert J, Austin A, Dart B, Bolding B, Maytubby P, Vogel JR, Stevenson BS. 2024. Wastewater Detection of Emerging Vector-Borne Diseases: West Nile Virus in Oklahoma. DOI: 10.2139/ssrn.4805820.

Lanciotti RS, Kerst AJ, Nasci RS, Godsey MS, Mitchell CJ, Savage HM, Komar N, Panella NA, Allen BC, Volpe KE, Davis BS, Roehrig JT. 2000. Rapid Detection of West Nile Virus from Human Clinical Specimens, Field-Collected Mosquitoes, and Avian Samples by a TaqMan Reverse Transcriptase-PCR Assay. Journal of Clinical Microbiology 38:4066–4071. DOI: 10.1128/jcm.38.11.4066-4071.2000.

McDonald E. 2021. Surveillance for West Nile Virus Disease — United States, 2009–2018. MMWR. Surveillance Summaries 70. DOI: 10.15585/mmwr.ss7001a1.

Monteiro S, Nunes F, Dosse M, Cangi Vaz N, Nhantumbo C, Juízo DL, Santos R. 2025. Environmental Surveillance of Vector-Borne Diseases in a Non-Sewered System: A Case Study in Mozambique. Environmental Science & Technology 59:3411–3421. DOI: 10.1021/acs.est.4c09860.

Monteiro S, Pimenta R, Nunes F, Cunha MV, Santos R. 2023. Wastewater-based surveillance for tracing the circulation of Dengue and Chikungunya viruses. :2023.10.30.23297765. DOI: 10.1101/2023.10.30.23297765.

Nebraska Department of Health and Human Services. 2025. West Nile Virus & Mosquito Surveillance Data. Available at https://dhhs.ne.gov/Pages/West-Nile-Virus-Data.aspx (accessed April 2, 2025).

Paz S. 2015. Climate change impacts on West Nile virus transmission in a global context. Philosophical Transactions of the Royal Society B: Biological Sciences. DOI: 10.1098/rstb.2013.0561.

Peccia J, Zulli A, Brackney DE, Grubaugh ND, Kaplan EH, Casanovas-Massana A, Ko AI, Malik AA, Wang D, Wang M, Warren JL, Weinberger DM, Arnold W, Omer SB. 2020. Measurement of SARS-CoV-2 RNA in wastewater tracks community infection dynamics. Nature Biotechnology 38:1164–1167. DOI: 10.1038/s41587-020-0684-z.

Roldan-Hernandez L, Van Oost C, Boehm A. 2024. Solid-liquid partitioning of Dengue, West Nile, Zika, Hepatitis A, Influenza A, and SARS-CoV-2 viruses in wastewater from across the United States. Environmental Science: Water Research & Technology. DOI: 10.1039/D4EW00225C.

Ronca SE, Ruff JC, Murray KO. 2021. A 20-year historical review of West Nile virus since its initial emergence in North America: Has West Nile virus become a neglected tropical disease? PLOS Neglected Tropical Diseases 15:e0009190. DOI: 10.1371/journal.pntd.0009190.

Santini M, Haberle S, Židovec-Lepej S, Savić V, Kusulja M, Papić N, Višković K, Župetić I, Savini G, Barbić L, Tabain I, Kutleša M, Krajinović V, Potočnik-Hunjadi T, Dvorski E, Butigan T, Kolaric-Sviben G, Stevanović V, Gorenec L, Grgić I, Glavač F, Mehmedović A, Listeš E, Vilibić-Čavlek T. 2022. Severe West Nile Virus Neuroinvasive Disease: Clinical Characteristics, Short- and Long-Term Outcomes. Pathogens 11:52. DOI: 10.3390/pathogens11010052.

Southern Nevada Health District. 2025. Mosquito Surveillance. Available at https://www.southernnevadahealthdistrict.org/programs/mosquito-surveillance/ (accessed March 30, 2025).

Topol A, Wolfe M, White B, Wigginton K, Boehm A. 2021a. High Throughput pre-analytical processing of wastewater settled solids for SARS-CoV-2 RNA analyses. protocols.io. DOI: dx.doi.org/10.17504/protocols.io.btyqnpvw.

Topol A, Wolfe M, Wigginton K, White B, Boehm A. 2021b. High Throughput RNA Extraction and PCR Inhibitor Removal of Settled Solids for Wastewater Surveillance of SARS-CoV-2 RNA. protocols.io. DOI: dx.doi.org/10.17504/protocols.io.btyrnpv6.

Vollans M, Day J, Cant S, Hood J, Kilpatrick AM, Kramer LD, Vaux A, Medlock J, Ward T, Paton RS. 2024. Modelling the temperature dependent extrinsic incubation period of West Nile Virus using Bayesian time delay models. Journal of Infection 89:106296. DOI: 10.1016/j.jinf.2024.106296.

Wolfe MK, Paulos AH, Zulli A, Duong D, Shelden B, White BJ, Boehm AB. 2024. Wastewater Detection of Emerging Arbovirus Infections: Case Study of Dengue in the United States. Environmental Science & Technology Letters 11:9–15. DOI: 10.1021/acs.estlett.3c00769.

World Health Organization. West Nile virus. Available at https://www.who.int/news-room/fact-sheets/detail/west-nile-virus (accessed March 30, 2025).

